# Dietary vitamin D intake and sun exposure are not associated with type 1 diabetic schoolchildren and adolescents: a first report in Algerian Sahara

**DOI:** 10.1101/2022.07.17.22276883

**Authors:** Slimane Brikhou, Wafa Nouari, Sofiane Bouazza, Chahrazed El Mezouar, Zakaria Benzian, Kheira Talha, Mourad Aribi

## Abstract

**Background:** A great number of children and adolescents worldwide suffer from physiological Vitamin D (VD) deficiency, which has been associated to the sun exposure and, consequently, to the risk of the development of various autoimmune diseases, including type 1 diabetes (T1D). However, the association of the disease with VD intake and sun exposure have yet to be explored.

**Materials and methods:** We conducted a food frequency questionnaire and 24-hour recall food survey, using “Ciqual table 2016” in 335 type 1 diabetic and age- and gender-matched healthy Algerian school children and teenager pupils from sunny Saharan and relatively less sunny Northern regions, aged between 5 and 19 years old.

**Results:** Both dietary VD intake and VD levels were similar in T1D patients when comparing between North and South regions (for the two comparisons, *p* > 0.05). Neither sun exposure, nor VD intake was associated with the disease (respectively, relative risk [RR] = 1.050, *p* = 0.680; RR = 1.082, *p* = 1.000. For Cochran and Mantel-Haenszel analysis; RR = 0.841, *p* = 0.862). VD intake showed a significant difference between diabetics and non-diabetics in sunny region (*p* = 0.022). Additionally, significant differences were highlighted between normal and T1D schoolboys (*p* = 0.038), and when comparing the two groups according to the dry areas (*p* = 0.016). Moreover, in contrast with the levels of circulating VD, which is decreased in T1D patients than in healthy controls, those of VD intake was significantly higher (*p* < 0.05), especially in male patients and in those with balanced diet, poor protein or carbohydrate consumption, a particular food intolerance, and a regular meal (*p* < 0.05), as well as in patients with a moderate or low consumption of cooked meals or steamed food (*p* < 0.01). Conversely, VD intake was markedly lower in type 1 diabetics than in controls regarding dry sunny region, including Adrar area, as well as in low fatty foods and eggs consumption (*p* < 0.05 for all comparisons). Nevertheless, relative risk of sun exposure and dietary vitamin D intake according to the WHO standard showed no significant association with T1D (common Mantel-Haenszel estimation, RR = 0.841, 95% CI 0.118-5.973, *p* > 0.05).

**Conclusions:** T1D seems to be not associated with VD intake and sun exposure in the Algerian Sahara region. Therefore, the consumption of VD in T1D patients from the Algerian Sahara would suspect that its association with the disease would be related to its synthesis alteration.

## 1. Introduction

T1D is an autoimmune disease, in which insulin-producing pancreatic β-cells are damaged progressively and selectively [1], and results from the interaction between genetic and environmental factors, including nutritional factors, like VD [2].

Involvement of VD in T1D pathogenesis has been suggested thanks to the observation about the difference in the disease incidence between North and South areas, suggesting it association with both low sun exposure and level of VD intake [3].

VD is represented as a micronutrient by two main anti-rachitic forms, including ergocalciferol (or VD_2_) and cholecalciferol (or VD_3_) [4]. VD_2_ is given from vegetables and fungi, and produced by irradiating plants and mushrooms with the ultraviolet B (UVB) rays [5,6], whereas VD_3_ has two main origins, *i*.*e*., dietary origin and provided essentially by fatty fish, liver, butter, cheese and egg yolk, etc., and endogenous origin, in which it is produced from photolyzed cholesterol by sun UVB rays [7]. Both VD_2_ and VD_3_ are absorbed to reach the circulation, tied to vitamin D-binding protein (VDBP), then it undergoes two main changes in liver to produce calcidiol then, in the kidneys, to generate the calcitriol [8–10].

VD poor diet, combined with insufficient exposure to the sunlight, causes VD deficiency at all life stages [11], and has been observed to be implicated in the development of T1D in childhood and adolescence [3]

In Algeria, the number of children under 15-year-old developing T1D is increasing, recording almost 3000 newly diagnosed patients with T1D every year [12]. Compared with other Mediterranean countries such as France, Greece and Italy, Algeria progressively evolved to reach 31.12 ± 3.60 (IC 95%, 27.90-34.23), between 2013 and 2017 [13].

Of note, although Algeria is characterized with a sunny climate around the year because of its location, there is, however, a notable blood VD deficiency in children and teenagers [14,15].

In this context, we have tried for the first time to check if dietary VD intake and normally sun exposure would be associated in young patients with recent onset T1D. Therefore, we assessed the VD intake in type 1 diabetic patients and age- and gender-matched healthy school children and teenagers from two Algerian regions according to the sun exposure levels, *i*.*e*., North with relative low sun exposure and Saharan with high relative sun exposure.

## 2. Population and methods

### 2.1 Study design and participants

The present study is a 24-hour recall food survey. The assessment of dietary VD intake was estimated using ciqual table 2016 [16]. Three hundred and thirty-five children and adolescents, including diabetics and non-diabetics aged between 5 and 19 years and age- and gender-matched, attending primary, middle and secondary schools, registered in screening and follow-up health school units (SFHSU) (Figure 1).

**Figure 1:**
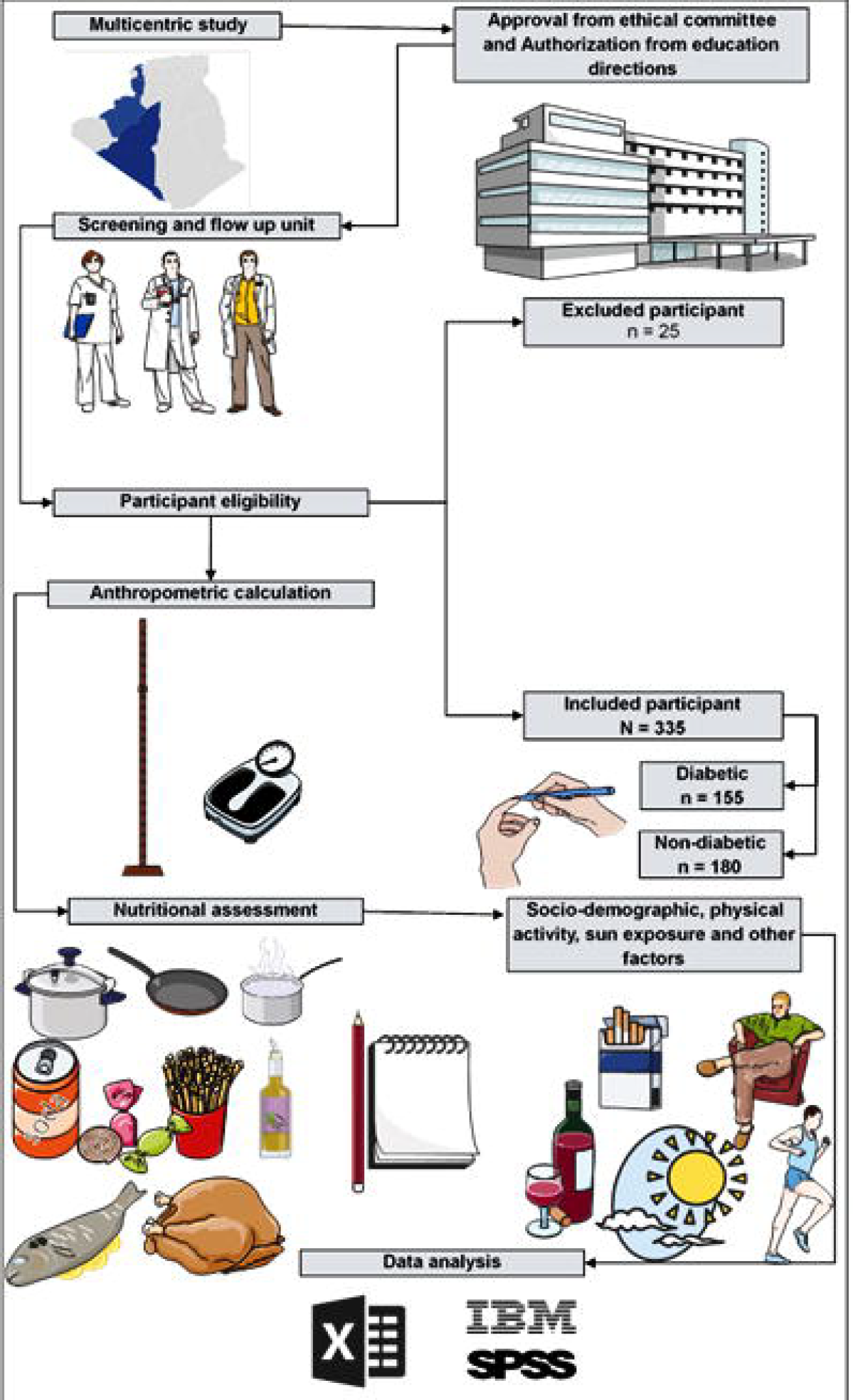
Summary and flowchart of the current study.

In each district, schools were randomly chosen according to their location in urban and rural areas, where all social classes coexist. In addition, at this age categories, education is compulsory in Algeria. Moreover, these schools are mixed and include almost the same number and structure of distribution of gender (boys and girls) and age groups, and with regard to the choice of age groups, this is for practical and physiological reasons. Furthermore, the selection of diabetic pupils was guided by the doctors of the SFHSU based on the WHO and EURODIAB diagnosis criteria [17].

### 2.2 Study geolocation and nutritional assessment

In the African continent Algeria is the largest country with an area of 2,381,741 km^2^ (919,595 Sq mile) and is estimated to have a population of around 44 million inhabitants. Approximately 70% of the population lives in the coastal region, the others live in the Sahara region mainly concentrated in oases, and almost 30% of Algerians are under 15 years of age [18].

This study was conducted in two areas of Algeria according to the intensity of sun exposure, including relative high sun exposure, *i*.*e*., Sahara (Naama, Bechar and Adrar), and relative low sun exposure, *i*.*e*., North of region of Algeria (Oran, Ain Temouchent, Sidi Bel-Abbès) (Figure 2). Diabetic pupils over 19 years and those who provided incomplete and erroneous information (especially the food quantity consumed) were excluded.

**Figure 2:**
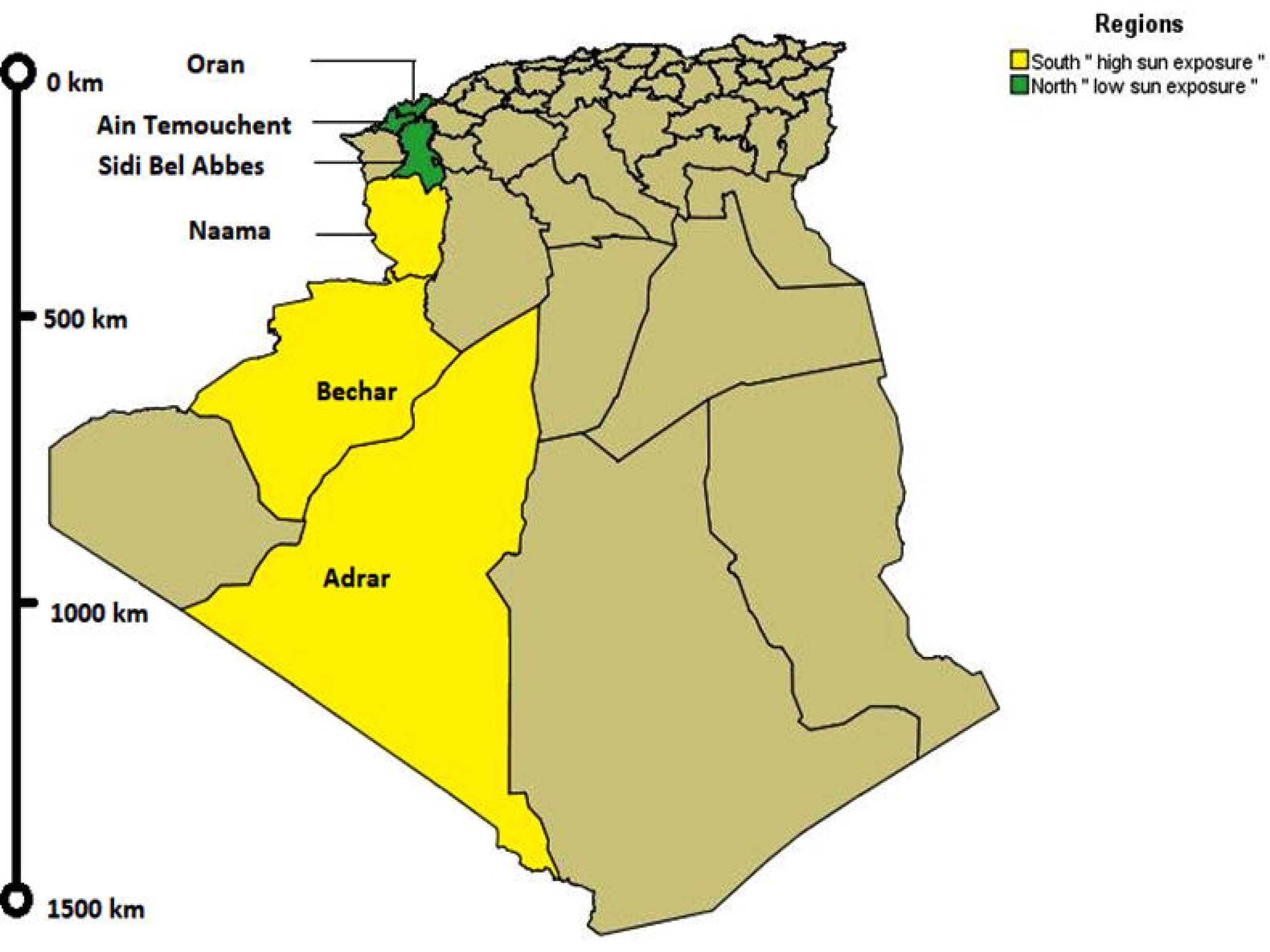
Geographical location of the study areas.

The questionnaire contains two sections, the first one includes personal information, socio-demographic, clinical and anthropometric parameters (age, gender, education level, height, weight, family history of diabetes, physical activity, eating and smoking habits). Skin phototypes were assessed according to Fitzpatrick’s classification. Weight and height measurement are obtained from the student clinical files. The second part of the questionnaire comprises two sections, a 24-hour recall that contains the dietary intake with details on all foods, water and beverages consumed during the previous day. The second one includes a food frequency questionnaire in which the informants specify the snack and certain type of food consumption, the cooking method, VD supplements, and foods rich in VD. Concerning the amount of the food, we used usual culinary instruments such as spoons, bowls, cups, plates, slice (slice of bread/cake), etc., and a food scale, moreover, each pupil was asked to precise even the portion was large, medium or small.

### 2.3 Body mass index

After obtaining data with CDC calculator [19], the sample was divided into four groups of body mass index (BMI) percentiles: underweight (< 5^th^ percentile), normal weight (5^th^ - 85^th^ percentile), overweight (85^th^ - 95^th^ percentile), and obese (≥ 95^th^ percentile) (Figure 3).

**Figure 3:**
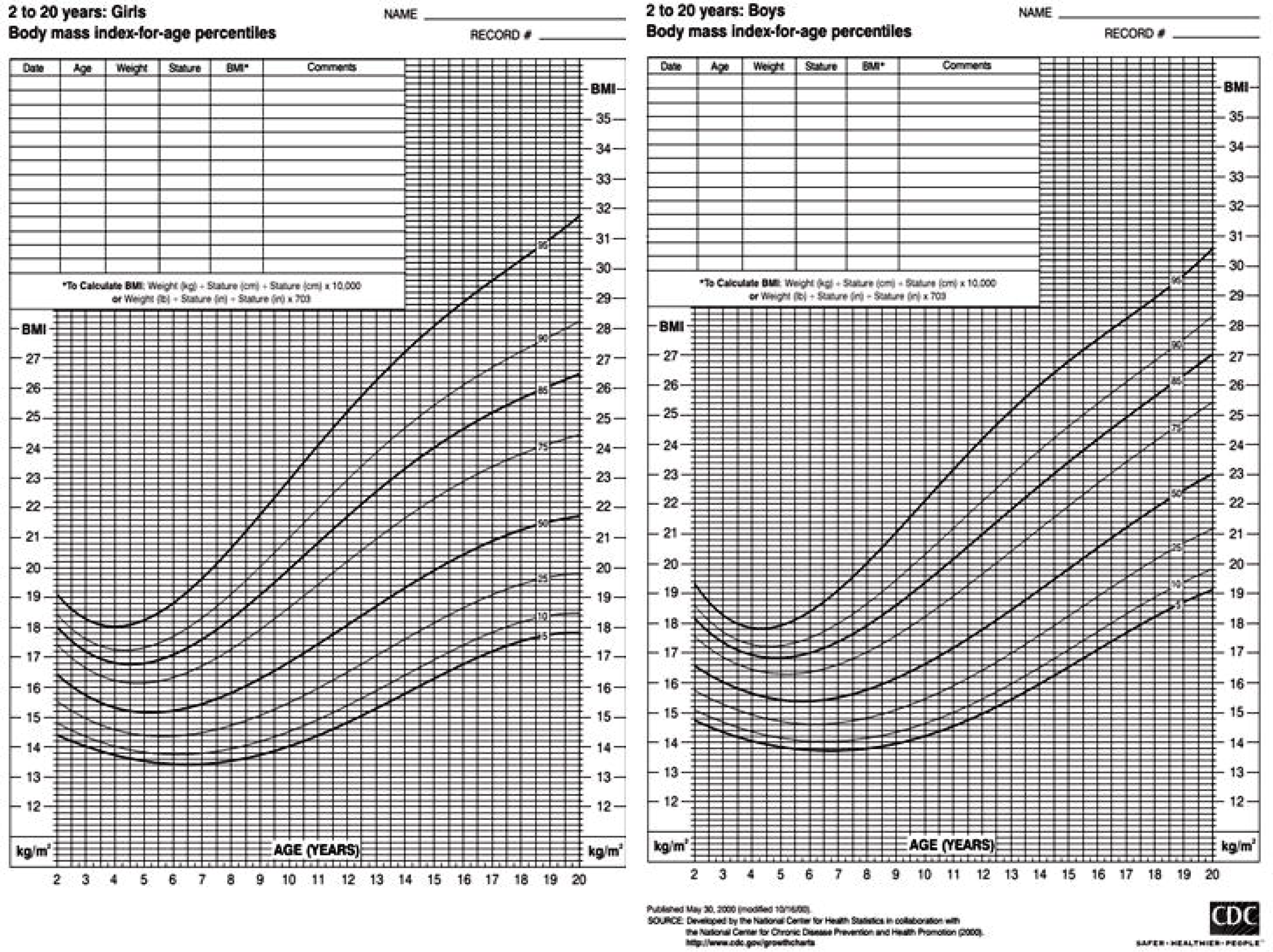
Percentile growth charts [19].

### 2.4 Basal VD

Basal circulating VD levels were recorded in all participants to check for any abnormality in serum vitamin D concentration prior to nutritional survey.

### 2.5 Ethical statement

The study was approved by the Institutional Review Board of Biology Department, Faculty of Life Sciences, University of Sidi-Bel-Abbès, Algeria, and authorized by the Direction of Education Authorities of each district, in collaboration with SFHSU Doctors. Students and even their parents were agreed to fill the questionnaire.

### 2.6 Statistical analyses

The collected data were represented as mean ± standard deviation, the statistical analyses was performed using IBM SPSS v 26 software. Estimation of the median was calculated at 95% confidence interval. Interquartile range was determined by the 25^th^ and 75^th^ percentiles. The comparison of VD intake, between groups, was done by *U* Mam Whitney test. The association between T1D and dietary VD intake was done using χ^2^ or Fisher exact test and the relative risk was estimated with 95% confidence interval. Cochran and common Mantel-Haenszel analysis was done to asses combined relative risk. Pearson correlation was used to verify the relation between dietary VD intake and latitude, sun exposure duration and body weight. The differences were considered as, significant when *p* < 0.05, highly significant if *p* < 0.01.

## 3. Results

The studied sample composed of T1D patients and controls. The comparison of VD intake expressed as microgram per day between them is mentioned in Table 1.

**Table 1.** Comparison of dietary VD intake (µg/d) in diabetic and non-diabetic pupils.

| Groups | Min. | Max. | Mean ±<br>SD | IQR | | Median | 95% CI | | *p*-value |
| --- | --- | --- | --- | --- | --- | --- | --- | --- | --- |
|  |  |  |  | 25<sup>th</sup> | 75<sup>th</sup> |  | LL | UL |  |
| ND | 0.03 | 20.25 | 1.48 ± 3.32 | 0.30 | 1.09 | 0.75 | 0.60 | 0.76 | 0.073 |
| T1D | 0.03 | 20.35 | 1.43 ± 2.94 | 0.30 | 1.44 | 0.80 | 0.76 | 0.98 | |

95% CI: confidence interval of the median, IQR: Interquartile range; LL: lower limit, ND: non-diabetics, SD: standard deviation, T1D: type 1 diabetics, UL: upper limit.

For the comparison between normal and T1D pupils, the overall population was divided into subgroups according to general characteristics, geographic localisation, eating habits, cooking methods, and foods rich in VD, as presented in Table 2.

**Table 2.** Comparison of dietary VD intake (µg/d) between diabetic and non-diabetics pupils.

| Variable |  | Diabetic<br>(n = 155) |  | Non-diabetics<br>(n = 180) |  | *p*-value |
| --- | --- | --- | --- | --- | --- | --- |
| | | n | $\bar{X} \pm \text{SD}$ | n | $\bar{X} \pm \text{SD}$ | |
| Socio-demographic | **Gender** |  |  |  |  |  |
| | Male | 68 | $1.92 \pm 2.43$ | 87 | $1.09 \pm 2.43$ | 0.038* |
| | Female | 87 | $1.06 \pm 1.83$ | 93 | $1.85 \pm 3.95$ | 0.456 |
|  | **Skin color** |  |  |  |  |  |
| | Black | 7 | $1.52 \pm 1.13$ | 5 | $0.45 \pm 0.21$ | 0.381 |
| | White | 55 | $1.41 \pm 3.20$ | 73 | $1.39 \pm 3.15$ | 0.061 |
| | Dark-brown | 92 | $1.45 \pm 2.93$ | 102 | $1.58 \pm 3.50$ | 0.519 |
|  | **Age** |  |  |  |  |  |
| | 5-10 | 28 | $0.91 \pm 0.60$ | 5 | $0.54 \pm 0.45$ | 0.268 |
| | 10-15 | 61 | $1.29 \pm 2.87$ | 92 | $1.50 \pm 3.34$ | 0.388 |
| | >15 | 66 | $1.83 \pm 3.58$ | 83 | $1.51 \pm 3.39$ | 0.185 |
|  | **BMI** |  |  |  |  |  |
| | Underweight | 9 | $0,66 \pm 0,59$ | 11 | $2,24 \pm 4,33$ | 0,824 |
| | Normal weight | 73 | $1,56 \pm 3,28$ | 116 | $1,51 \pm 3,38$ | 0,118 |
| | Overweight | 13 | $1,87 \pm 3,74$ | 5 | $0,49 \pm 0,32$ | 0,289 |
| | Obese | 18 | $1,00 \pm 0,79$ | 7 | $0,49 \pm 0,28$ | 0,074 |
|  | **Study Level** |  |  |  |  |  |
| | Primary | 36 | $0.90 \pm 0.75$ | 33 | $1.14 \pm 2.82$ | 0.134 |
| | Middle | 66 | $1.26 \pm 2.88$ | 71 | $1.47 \pm 3.31$ | 0.412 |
| | Secondary | 53 | $2.09 \pm 3.91$ | 76 | $1.63 \pm 3.55$ | 0.157 |
|  | **Sport** |  |  |  |  |  |
| | No | 33 | $0.23 \pm 0.61$ | 16 | $0.59 \pm 0.45$ | 0.344 |
| | Yes | 122 | $1.59 \pm 3.26$ | 164 | $1.87 \pm 3.47$ | 0.091 |
| **Geographical location** | **Regions** |  |  |  |  |  |
| | Region with relative low sun exposure “North” | 74 | $1.61 \pm 3.89$ | 90 | $1.56 \pm 1.08$ | 0.906 |
| | Ain Témouchent | 30 | $1.55 \pm 4.23$ | 30 | $1.86 \pm 4.36$ | 0.560 |
| | Sidi Bel Abbès | 30 | $2.11 \pm 4.34$ | 30 | $1.64 \pm 3.03$ | 0.823 |
| | Oran | 14 | $0.60 \pm 0.60$ | 30 | $1.12 \pm 2.74$ | 0.878 |
| | Region with relative high sun exposure “South” | 81 | $1.28 \pm 1.80$ | 90 | $1.39 \pm 1.14$ | 0.022* |
| | Naama | 21 | $1.77 \pm 3.30$ | 30 | $1.85 \pm 2.98$ | 0.946 |
| | Bechar | 30 | $1.25 \pm 0.65$ | 30 | $0.63 \pm 0.51$ | 0.018* |
| | Adrar | 30 | $1.19 \pm 0.93$ | 30 | $1.76 \pm 4.94$ | 0.027* |
|  | **Climate** |  |  |  |  |  |
| | Dry | 83 | $1.35 \pm 1.87$ | 90 | $1.39 \pm 3.13$ | 0.016* |
| | Humide | 72 | $1.53 \pm 3.88$ | 90 | $1.56 \pm 3.48$ | 0.777 |
| **Diet habits** | **balanced diet** |  |  |  |  |  |
| | No | 56 | $1.01 \pm 1.32$ | 30 | $1.25 \pm 2.80$ | 0.647 |
| | Yes | 99 | $1.65 \pm 3.47$ | 150 | $1.53 \pm 3.43$ | 0.036* |
|  | **Rich in carbohydrates** |  |  |  |  |  |
| | No | 118 | $1.58 \pm 3.27$ | 160 | $1.49 \pm 3.33$ | 0.016* |
| | Yes | 37 | $0.99 \pm 1.50$ | 20 | $1.42 \pm 3.38$ | 0.787 |
|  | **Rich in protein** |  |  |  |  |  |
| | No | 116 | $1.57 \pm 3.29$ | 166 | $1.56 \pm 3.43$ | 0.034* |
| | Yes | 39 | $0.84 \pm 1.48$ | 14 | $0.52 \pm 0.40$ | 0.456 |
|  | **Rich in lipids** |  |  |  |  |  |
| | No | 132 | $1.50 \pm 3.08$ | 167 | $1.45 \pm 3.26$ | 0.090 |
| | Yes | 23 | $0.84 \pm 0.70$ | 13 | $1.89 \pm 4.11$ | 0.519 |
|  | **Intolerance to a particular food** |  |  |  |  |  |
| | No | 135 | $1.32 \pm 2.51$ | 153 | $1.07 \pm 2.32$ | 0.016* |
| | Yes | 20 | $2.41 \pm 5.43$ | 27 | $3.96 \pm 6.24$ | 0.431 |
|  | **The habit of skipping a meal** |  |  |  |  |  |
| | No | 56 | $1.81 \pm 3.43$ | 68 | $1.12 \pm 2.25$ | 0.010* |
| | Yes | 98 | $1.18 \pm 2.56$ | 112 | $1.68 \pm 3.76$ | 0.857 |
| Cooking method | **Grilled** |  |  |  |  |  |
| | No | 148 | $1.32 \pm 2.76$ | 165 | $1.31 \pm 2.83$ | 0.100 |
| | Yes | 7 | $3.39 \pm 5.24$ | 15 | $3.06 \pm 6.14$ | 0.207 |
|  | **Fried** |  |  |  |  |  |
| | No | 87 | $1.26 \pm 2.51$ | 69 | $1.63 \pm 3.37$ | 0.679 |
| | Yes | 68 | $1.65 \pm 3.41$ | 111 | $1.41 \pm 3.31$ | 0.086 |
|  | **Boiled** |  |  |  |  |  |
| | No | 115 | $1.40 \pm 2.69$ | 114 | $1.06 \pm 2.38$ | 0.009** |
| | Yes | 40 | $1.52 \pm 3.67$ | 66 | $2.13 \pm 4.33$ | 0.759 |
|  | **Steamed** |  |  |  |  |  |
| | No | 129 | $1.58 \pm 3.26$ | 108 | $1.31 \pm 3.13$ | 0.005** |
| | Yes | 26 | $1.18 \pm 2.13$ | 72 | $1.71 \pm 3.57$ | 0.356 |
|  | **In Sauce** |  |  |  |  |  |
| | No | 21 | $2.95 \pm 5.70$ | 19 | $0.78 \pm 0.62$ | 0.257 |
| | Yes | 134 | $1.18 \pm 2.13$ | 161 | $1.59 \pm 3.55$ | 0.134 |
| Foods rich in VD | **Fish** |  |  |  |  |  |
| | No | 88 | $0.94 \pm 0.74$ | 89 | $0.95 \pm 1.91$ | 0.074 |
| | Yes | 66 | $2.07 \pm 4.32$ | 91 | $1.95 \pm 4.14$ | 0.373 |
|  | **Fats** |  |  |  |  |  |
| | No | 111 | $1.16 \pm 1.66$ | 131 | $1.40 \pm 3.14$ | 0.049* |
| | Yes | 43 | $2.15 \pm 4.90$ | 49 | $1.66 \pm 3.72$ | 0.848 |
|  | **Eggs** |  |  |  |  |  |
| | No | 61 | $1.55 \pm 3.62$ | 46 | $1.26 \pm 3.39$ | 0.633 |
| | Yes | 93 | $1.36 \pm 2.46$ | 134 | $1.56 \pm 3.31$ | 0.042* |
|  | **Cheese** |  |  |  |  |  |
|  | No | 40 | 1.62 ± 2.73 | 31 | 0.92 ± 1.35 | 0.178 |
|  | Yes | 114 | 1.38 ± 3.01 | 149 | 1.59 ± 3.56 | 0.204 |
|  | **Cereals** |  |  |  |  |  |
|  | No | 54 | 1.46 ± 3.25 | 68 | 1.15 ± 2.56 | 0.132 |
|  | Yes | 100 | 1.42 ± 2.78 | 112 | 1.71 ± 3.74 | 0.320 |

\* $p < 0.05$ , \*\* $p < 0.01$ . BMI: body mass index classes.

As depicted in Figure 4, dietary VD intake percentiles were not similar in both groups. In fact, the difference was observed upper the 35^th^ percentile and was clearly evident in subjects having more than 90^th^ percentile. Such a result indicates that less than 5% of all subjects have a good dietary VD intake.

**Figure 4:**
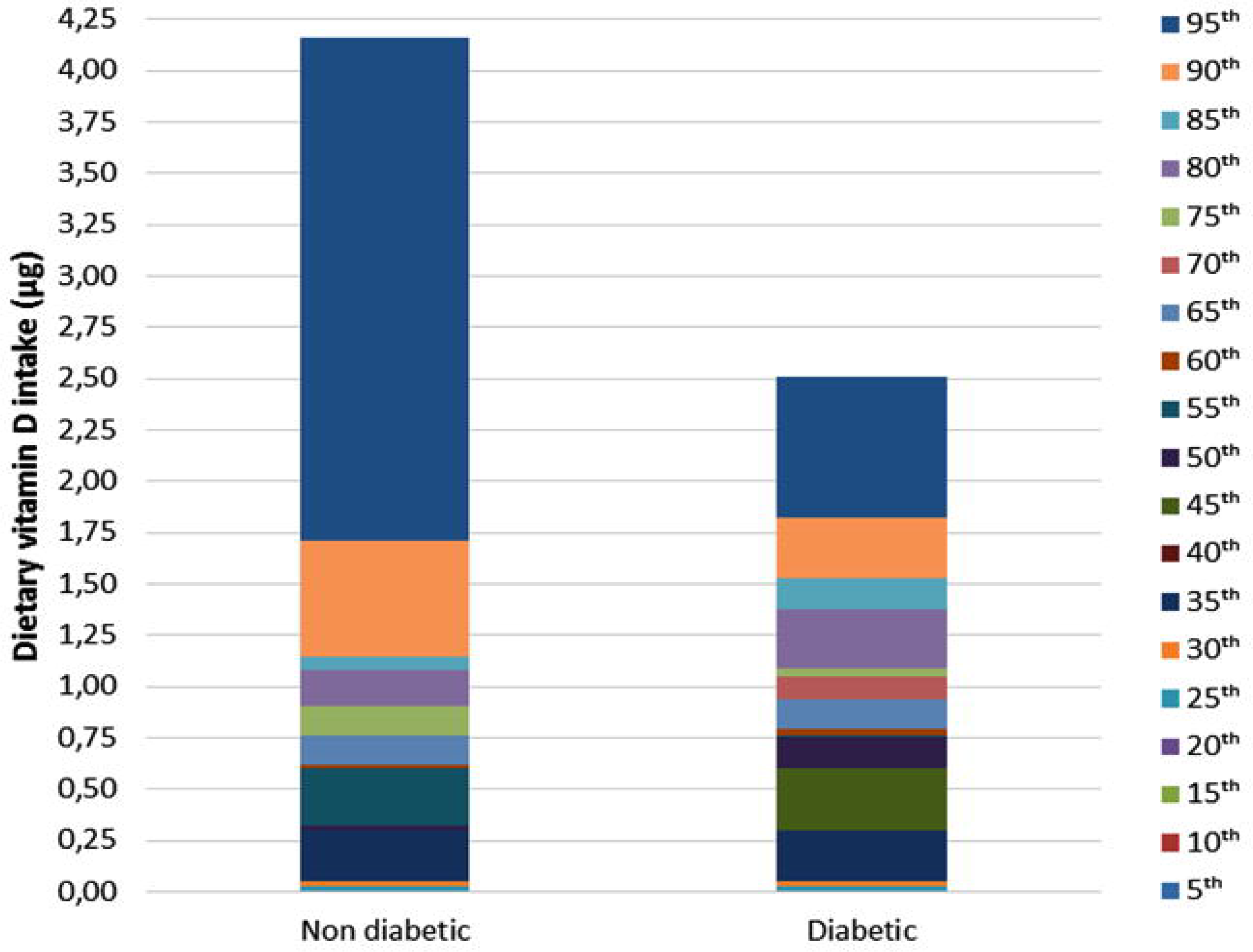
Dietary vitamin D intake percentiles in both groups.

In contrast to the basal circulating VD levels, according to the WHO standard, that were decreased in T1D patients than in healthy controls, regardless of geographic locality with respect to sun exposure (*p* < 0.05) (data not shown), our results did not show a significant difference between VD intake distribution in diabetic and non-diabetic pupils (Table 1).

As shown in Table 2, skin, age, educational levels, and sport practicing, did not make any significant difference between T1D patients and healthy controls (for all comparisons, *p* > 0.05). Additionally, we observed that dietary VD intake medians was significantly increased in T1D patients than in ND subjects, according to the region with relative high sun exposure (*p* = 0.018).

There was a moderate positive and highly significant correlation (n = 61, Spearman’s Rho = 0.372, *p* = 0.003) between VD intake and body weight gain in healthy population, conversely to diabetics (Table 3). On other hand, no significant correlation was observed between dietary VD intake, body weight lost, and latitude as well as sun exposure duration for diabetics and non-diabetics pupils (Figure 5).

**Table 3.** Bivariate correlation of latitude, sun exposure duration and body weight with vitamin D intake.

| **Variable** | **Non-diabetics** |  |  | **Type 1 diabetics** |  |  |
| --- | --- | --- | --- | --- | --- | --- |
|  | **n** | ***r*** | ***p*** | **n** | ***r*** | ***p*** |
| **Latitude** | 180 | -0.062 | 0.409 | 155 | -0.035 | 0.667 |
| **Sun exposure duration** | 180 | 0.000 | 0.995 | 155 | -0.092 | 0.256 |
| **Body weight lost** | 54 | -0.072 | 0.605 | 49 | -0.031 | 0.833 |
| **Body weight gain** | 61 | 0.372** | 0.003 | 65 | -0.017 | 0.893 |

\*\* $p < 0.01$ .

**Figure 5.**
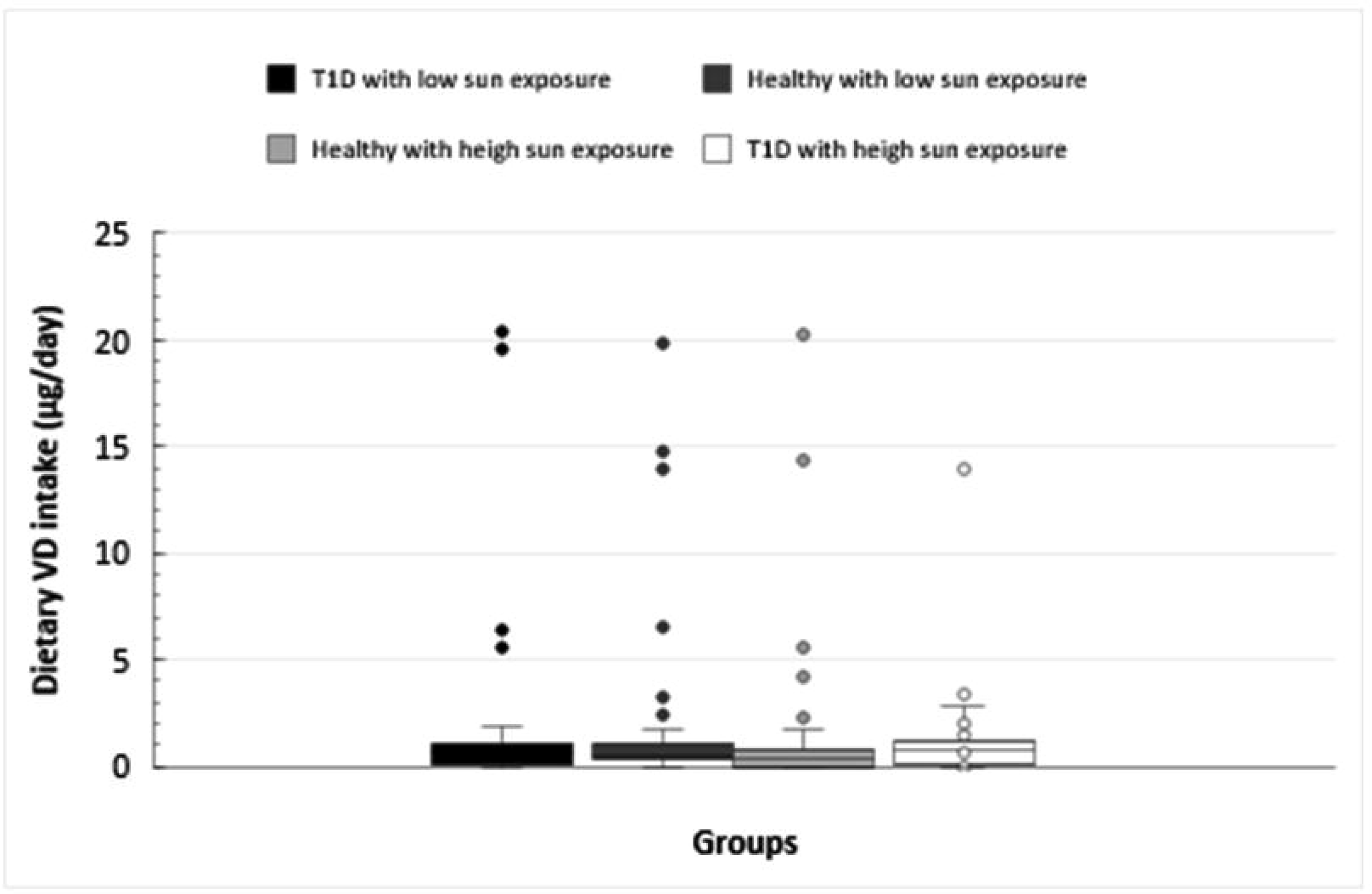
Levels of vitamin D intake in type 1 diabetes and healthy controls according to sun exposure.

Comparisons according to the regions, a significant difference in dietary VD intake was shown in Southwest of Algeria (*p* = 0.022), especially in Adrar (*p* = 0.027), and Bechar (*p* = 0.018); while such a difference remains identical in the Northern region. In addition, except to humidity, there was a significant difference in VD intake between normal and T1D schoolboys (*p* = 0.038); while schoolgirls showed a similar VD intake (*p* = 0.456). Furthermore, a significant difference in the dry areas was observed (*p* = 0.016) more specifically in Adrar and Bechar regions.

As for diet habits, schoolchildren having a balanced diet, differ significantly in dietary VD intake. Learners who did not consume a diet rich in carbohydrate or protein, a marked difference was observed in term of dietary VD intake (*p* = 0.030). Additionally, for pupils who did not declare an intolerance to a specific food such as fish, fat, eggs, an important difference was observed (*p* = 0.016). Regarding the meal skip habit, a significant difference was clearly indicated for those who did not skip their meals (*p* = 0.010). Taking in consideration the cooking method, a high significant difference, was observed in subjects who did not use the boiled method (*p* = 0.009). For the subjects who did not use the streamed method, a high significant difference was observed (*p* = 0.005). As for the other cooked methods (grilled, fried, and in sauce), no significant difference was shown (*p* > 0.05). The results indicate that consuming the food rich in VD such as fats displays a difference in dietary VD intake (*p* = 0.049), contrary to fish, cheese and cereals, no significant difference was observed. For the informants who consumed the eggs, a significant difference was noted (*p* = 0.042).

Finally, no association was observed for common Mantel-Haenszel estimation when considering both sun exposure and dietary VD intake according to the WHO standard (relative sun exposure; RR, 1.050, 95% CI 0.833-1.323, dietary VD intake; RR, 1.082, 95% CI 0.403-2.902, MH; RR, 0.841, 95% CI 0.833-1.323, 0.118-5.973, *p* =0.862) (Table 4).

**Table 4.** Relative risk of sun exposure and dietary vitamin D intake according to the WHO standard.

| **Factors** | **RR** | **95% CI** |  | ***p*-value** |
| --- | --- | --- | --- | --- |
|  |  | **LL** | **UL** |  |
| **Relative sun exposure** | 1.050 | 0.833 | 1.323 | 0.680 |
| **Dietary VD intake** | 1.082 | 0.403 | 2.902 | 1.000 |
| **MH** | 0.841 | 0.118 | 5.973 | 0.862 |

95% CI: 95 % confidence interval, LL: lower limit, MH: common Mantel-Haenszel estimation, RR: relative risk, UL: upper limit.

## 4. Discussion

VD can be available from sunlight, and it varies related to endogenous factors such as : skin pigmentation, genetics, physiology (bioavailability, bioequivalence and safety), adiposity, bile acid secretion and exogenous factors: socio-demographic (age, income, race/ethnicity, gender), physical inactivity, nutrition (food VD intake (food sources (food rich in VD, competing foods (cholesterol, tocopherol, phytosterols [20]), diet habits (cooking method, including skipping a meal, dairy and animal product exclusion (lactose intolerance, cow’s milk food allergy [21], ovo-vegetarianism, veganism), VD supplementation and fortification, sun exposure (latitude, season, outdoor physical activity, use of sunscreens, cultural differences in dress, tanning bed), drugs (anticonvulsants (phenobarbital, dilantin, tegretol), bisphosphonates [22], ketoconazole, thiazides [23] and toxic habits including smoking. The classical role of VD to promote the human growth is very important in childhood and adolescence, in which the individual spends considerable time in education and school activities such as sport. In Algeria, studies about VD status in children and adolescents are scarce. According to our knowledge, the conducted research is the first one performed in Algeria that reports on the dietary VD intake in schoolchildren and adolescents, in the largely urban and rural areas of many districts. Through this study, we tried to study the difference in VD intake in food consumed by healthy and patient pupils with T1D.

### 4.1 VD deficiency

In 2005, a national survey, named Transition and Health Impact in North Africa (TAHINA) was carried out by the National Institute of Public Health of Algiers (INSP) in collaboration with the European Union in 16 districts, showed that the daily food consumption did not comply with international health recommendations [24], thus, the present study found that the reported food intake of VD was low. This situation may be explained by the low socioeconomic status and the consumption of VD poor diet [25,26]. People may reliant on diet to satisfy their VD requirements, but according to some reports, a high rate of VD insufficiency in Algerian children and teenagers was noticed [15,27], which are consistent with results obtained in other regions in the world such as, Spain, Greece, Africa [28,29]. In fact, the consumption of food rich in VD is little or almost non-existent and even fortified foods are negligible, for instance, the milk is generally not fortified with VD, unlike in Western countries [30], such as Finland [31], Germany, Ireland, the Netherlands, the United Kingdom, and the United States, in which higher intakes are remarked [32,33]. In our sample, cheese did not make a significant difference between the normal and diabetic pupils. Although Algeria is one of the major consumer countries as regards the milk and dairy products, but this consumption is primarily a source of calcium and protein [34].

### 4.2 Regions

According to O’Mahony, et al., 2011, dietary VD intake differ from region to another. Unlike Mediterranean countries, intakes are higher in Scandinavian region [35]. This lack of VD in foods may also be related to the availability and bioavailability of food containing VD [36]. In Japan, Norway, United Kingdom, and Spain the major sources of dietary VD intake are fish [36], while in Algerian Sahara, fresh fish arrive with difficulty and irregularly, due to the long distance between the fishing port and this region, moreover the distribution of small and very distant agglomerations, the high cost in winter, and the rapid deterioration in quality, especially by the heat in summer make this product rare and expensive.

Fat are also the most common food sources in Norway, that why VD status is adequate in a large part of the population [37,38]. Comparing with our sample, there was no difference in VD intake in pupils eating diet rich in lipids, so we hypothesise that in these diets the VD intake was very low.

### 4.3 Eating habits

Dietary intake habits may differ from country to another which make region and population-specific research of VD important [39]. In the United Kingdom, cereal and cereal products contribute significantly to average daily VD intakes in adults [40], whereas, our study showed that patients and healthy learners who did not consume a diet rich in carbohydrate or protein differ in VD intake. In Japan, eggs are the second major sources of dietary VD intake after fish [41], while, in patients and healthy pupils who did not consume a diet rich in fat were markedly different in VD intake. On top of that, there were certain foods that could not be tolerated by the diabetics and controls, such as milk, eggs, fish, etc., but, in fact, even these pupils had no intolerance or allergy to particular food, a significant difference was observed. For diabetic and healthy schoolchildren and adolescents who had a balanced diet, VD intake differed significantly.

Skipping meal as breakfast or lunch among adolescents and children worldwide, becomes a public health issue, that leads to insufficient intake of essential nutrients such as VD [42–44]. For our sample the significant difference in VD intake for those who did not skip any meal, may clarify that their diet was insufficient in VD intake, since lunch and breakfast are important sources to furnish this nutrient. For the cooking methods, previous studies revealed that cooking may affect VD retention in diet since VD is a micronutrient which is sensitive to cooking, and its loss is generally higher for longer cooking process or higher heating temperature [45]. VD intake was significantly different between the two groups except boiling and streaming methods which are not commonly used in Algerian kitchen.

### 4.4 Gender

From our findings, it appears that in both genders VD dietary intake is low, with a significant difference between normal and T1D schoolboys. Previous studies found that males had higher intakes of VD [46,47]. In Ireland, Black et *al*., 2014 [48] found that teenage boys had the highest intakes (2.7 µg/d), which were significantly higher than in teenage girls (1.9 µg/d) with no significant difference between boys and girls in users of VD-containing supplements. According to the Canadian Community Health Survey (CCHS), daily mean of VD intakes were 7.3 and 5.4 µg in 9-18-year-old boys and girls, respectively Vatanparast, et al., 2010, Murphy et *al*., compared milk intake which is the main source of VD in USA, and revealed sex differences [49].

### 4.5 Skin

For all researchers, the major and the essential source of VD is the skin synthesis [50], [51], [52], which represents 90 % of the individual’s needs in VD [53], [54], within 20 minutes, it can be obtained at least 10 000 (IU), while nutrients can only provide 40-400 IU [55], *i*.*e*., less than 10% [56] and from this, dietary sources represent small amount. But despite this situation, dietary intake may be remained important, especially in areas with limited sun exposure.

In Norway, skin with little pigment makes VD status from sunlight adequate, contrary to Algerian population and basing on data from countries with similar climates and the same human composition, the VD status was low [38]. The present study revealed that no significant difference in VD intake between controls and patients.

### 4.6 BMI

VD intake classed by BMI percentile did not differed for each group between normal and T1D pupils. Moore et *al*., (2014) reported that 15.2% were overweight (≥85^th^ to <95^th^ BMI percentile) and 19.1% were obese (BMI ≥ 95^th^ percentile) but did not give match details about VD intake in each group. It is known that increased adiposity lead to lower serum 25OHD levels [57]. Nonetheless, our study showed that VD intake was higher in obese persons compared with their leaner counterparts, and generally we can classify them as follow: underweight pupils (9.3%), normal weight (75.5%), overweight (7.2%), and Obese (8.1%). So, we should promote VD intake of dietary to these categories, since all of them are below the recommendation,

### 4.7 Diet survey

It is common that foods eaten less than once or twice a week may not be captured in one day dietary record, in addition to subject recall errors and underreporting [58]. Nonetheless, large group is sufficient to report mean usual intake using single 24-hour recall [50]. For these reasons, this survey was supplemented with a food frequency questionnaire [59].

### 4.8 Limitations

One of limitations is the uncertainties surrounding the Dietary Reference Intake (DRI) values for VD which leads to an inability to characterize and integrate sun exposure with dietary intake recommendations as much as may be appropriate [57]. Nevertheless, this was a food VD intake assessment, and cannot be compared to DRI because dietary supplementation was not recorded to estimate total VD intake.

## 5. Conclusions and future prospects

In the current study, although the basal circulating VD levels were decreased in T1D patients than in healthy controls, our results did not show an alteration in VD intake and sun exposure in T1D patients.

However, even then we would like to recommend VD supplementation for children and adolescents, especially in schools which are good and favourable places to implement many nutritional strategies, such as a VD campaign by vaccination, or a focus in school canteen programs in which we encourage pupils to eat foods rich in VD. This remains as an alternative, but not a definitive strategy, because alteration in VD levels may be due to an abnormality of enzyme bioconversion to the VD active, which, consequently, leads to its usually immunomodulatory effect.

## Data Availability

All data produced in the present work are contained in the manuscript

## Acknowledgments

First of all, the authors would like to thank the Algerian Ministry of Higher Education and Scientific Research, as well as the General Direction of Scientific Research and Technological Development (DGRSDT) for the support of this work. They also express their deep thanks to all the learners and their parents who contributed to this study, as well as the Educational Institutions for their valuable help.

